# Beyond malaria prevention: sulfadoxine-pyrimethamine treatment in pregnancy selectively remodels the maternal gut microbiome to increase gestational weight gain and improve birthweight

**DOI:** 10.64898/2026.05.03.26352319

**Authors:** Andreea Waltmann, Sydney M. Puerto-Meredith, Jobiba Chinkhumba, Enala Mzembe, Michael Kayange, Ian Carroll, Jeffrey Roach, Don P. Mathanga, Julie R. Gutman, Jonathan J. Juliano

## Abstract

Intermittent preventive treatment in pregnancy (IPTp) with sulfadoxine-pyrimethamine (SP), an antifolate drug with antimalarial and antibiotic activity, reproducibly improves birthweight across sub-Saharan Africa and the Western Pacific. This clinical protection is independent of SP’s original malaria indication: it is not diminished by widespread antimalarial resistance or reduced transmission, and SP outperforms more potent non-antibiotic antimalarials (e.g., dihydroartemisinin-piperaquine, DP) for fetal growth. The biological mechanism is unexplained. We previously showed that gestational weight gain (GWG) is a significant component of this mechanism and mediates two-thirds of SP’s overall birthweight benefit (NCT03009526). In the first longitudinal characterization of antifolate antibiotic effects on the pregnant gut microbiome, we show that ∼45% of SP’s GWG advantage over DP is explained by gut microbial changes consistent with its pharmacology. Microbiome-mediated GWG coincided with 126g higher birthweight in SP but not DP recipients (95%CI 22.6-229.3g; p=0.019). Relative to DP, SP suppressed gastrointestinal pathobionts and enriched anaerobic commensals with recognized roles in mucosal immunity and host metabolism, a microbiome-sparing pattern distinct from conventional antibiotic-associated dysbiosis.

## Main

Fetal growth *in utero* shapes lifelong health. Yet growth faltering in the womb remains a leading cause of stillbirth, and infant morbidity and mortality^1^. Worldwide, 22 million babies (∼1 in 5) are born at term but small-for-gestational age (SGA, <10^th^ percentile) and 8-10 million of them are small enough to be counted as low birthweight (<2,500 grams)^2^. While this burden is disproportionately concentrated in low-to-middle-income countries (LMICs)^2^, available evidence demonstrates that this is a global challenge. Fetuses of mothers living under favorable nutritional and health conditions across different geographical contexts showed similar growth trajectories^3^. Globally, maternal undernutrition and recurrent infections are two common preventable drivers of low birthweight^4^.

Few antenatal interventions implemented at scale have consistently improved birthweight, pointing to unaddressed microbial-nutritional pathways that impair fetal growth. Intermittent preventive treatment in pregnancy (IPTp) with monthly sulfadoxine-pyrimethamine (SP; 1500 mg sulfadoxine and 75 mg pyrimethamine) is a notable exception. It was introduced more than three decades ago into antenatal care programs in sub-Saharan Africa and later Papua New Guinea to reduce the risk of low birthweight due to maternal exposure to *Plasmodium falciparum* malaria in pregnancy ^5, 6^. The World Health Organization recommends at least 3 monthly doses of IPTp-SP starting in the second trimester to all pregnant women in *Plasmodium falciparum* malaria endemic regions^7^. In 2023, an estimated 13 million pregnant women in sub-Saharan Africa (44% of all pregnancies in 34 countries conducting IPTp) received ≥3 doses of IPTp-SP^7^.

SP is a broad-spectrum antifolate antibiotic and antiparasitic^8^. Antimicrobial resistance to SP is now widespread^5^, compromising SP’s antimalarial efficacy^9^, including in Malawi^9, 10^. Paradoxically, SP’s birthweight benefit has not diminished^5^. A meta-analysis spanning >100 studies and ∼150,000 pregnancies underscores this persistent effect^5^, which is not explained by prevention of preterm birth and exceeds the benefit of non-antibiotic antimalarials such as dihydroartemisinin-piperaquine (DP)^6^. These observations point to malaria-independent mechanisms through which SP improves fetal growth, that if identified could generate new knowledge of fetal growth biology with global clinical relevance and inform non-antibiotic interventions.

Why a drug introduced for malaria prevention retains a reproducible birthweight benefit after its antimalarial effect has been weakened remains an important unresolved question in maternal-child health. In low- and middle-income settings, where the major modifiable drivers of low birthweight often cluster around infection and undernutrition^4^, this pattern raises the possibility that SP addresses a shared microbial-nutritional pathway.

In this Malawian randomized controlled trial of IPTp-SP versus DP, we previously showed that increased gestational weight gain (GWG) mediates SP’s birthweight benefit^11^. GWG is a known determinant of fetal growth^12, 13^. Our findings have since been replicated in a large meta-analysis^14^. Because antibiotics can shift gut microbiota in ways that influence weight gain and metabolism in non-pregnant contexts^15^, we hypothesized that SP’s long-acting antimicrobial activity remodels the maternal gut microbiome to support GWG and fetal growth. This idea is consistent with SP’s well-described broad-spectrum antimicrobial mode of action and long half-life^16, 17^, including its absorption through the gut.

## Results

Baseline characteristics of the ancillary microbiome cohort by randomized group are shown in **Extended Data Table 1**.

### Samples and sequencing

We profiled 245 longitudinal stool samples from 91 women (46 SP, 45 DP) by 16S rRNA gene V4 amplicon sequencing in technical duplicate (490 total microbiomes). After quality filtering at a rarefaction depth of 51,000 reads, 243 microbiomes were retained for analysis (119 SP, 124 DP; mean 2.7 samples per woman). Sequencing quality metrics and replicate concordance are detailed in **Supplementary Note 1**, and **Supplementary Fig. 1 and 2**. Demultiplexed sequencing data is available in the NCBI Sequence Read Archive (SRA) and can be accessed via accession number PRJNA1416942.

### Global diversity changes after IPTp-SP relative to IPTp-DP

Compared to IPTp-DP, IPTp-SP did not alter global α- or β-diversity (**Supplementary Fig. 3**). Exploratory compositional analysis of 929 amplicon sequence variants (SVs) with the Gneiss tool over the course of treatment (n post-baseline microbiomes=173 microbiomes, SP=81, DP=92) revealed detectable changes in bacterial composition in response to SP relative to DP (**Extended Data Fig. 1, Supplementary Table 1**), motivating formal differential abundance analysis.

### Differential abundance analysis

After sparsity filtering (zero abundance in ≥80% of samples in each treatment group independently), 350 of 929 SVs were retained; following exclusion of 21 SVs differentially abundant at the pre-treatment baseline visit (p<0.05), 329 SVs were taken forward for differential abundance testing between study arms (**Supplementary Table 2**). Two independent statistical frameworks (limma-voom and generalized linear mixed models) were used, with good concordance in direction and magnitude of effect sizes (**Supplementary Fig. 4**).

We assessed differences in relative abundance of SVs across the entire IPTp period, in samples collected after ≥2 and after ≥3 IPTp doses. In total, 84 SVs (53 unique taxa) were found to be differentially abundant in SP vs. DP microbiomes (p<0.1, **Fig. 1**, **Supplementary Table 3**). Of these, 57 SVs (corresponding to 31 unique taxa) were enriched with SP vs. DP; 27 SVs (corresponding to 15 unique taxa) were reduced with SP vs. DP (**Fig. 1B, 1C**). Seven taxa contributed SVs to both the enriched and reduced directions (i.e., several SVs mapped with the same taxonomy), suggesting strain-level or niche-level heterogeneity. Several SVs showed consistent differences from the first dose onward while others emerged after ≥2 or ≥3 doses (Fig. 1B, C). Taxonomic resolution was improved by consulting additional reference databases (NCBI or Integrated Microbial Genomes, IMG): of the 84 SVs, 52 (61.9%) were classified to species level, 18 (21.4%) to genus, and 14 (16.7%) to family or order (**Supplementary Table 3**).

**Fig. 1.**
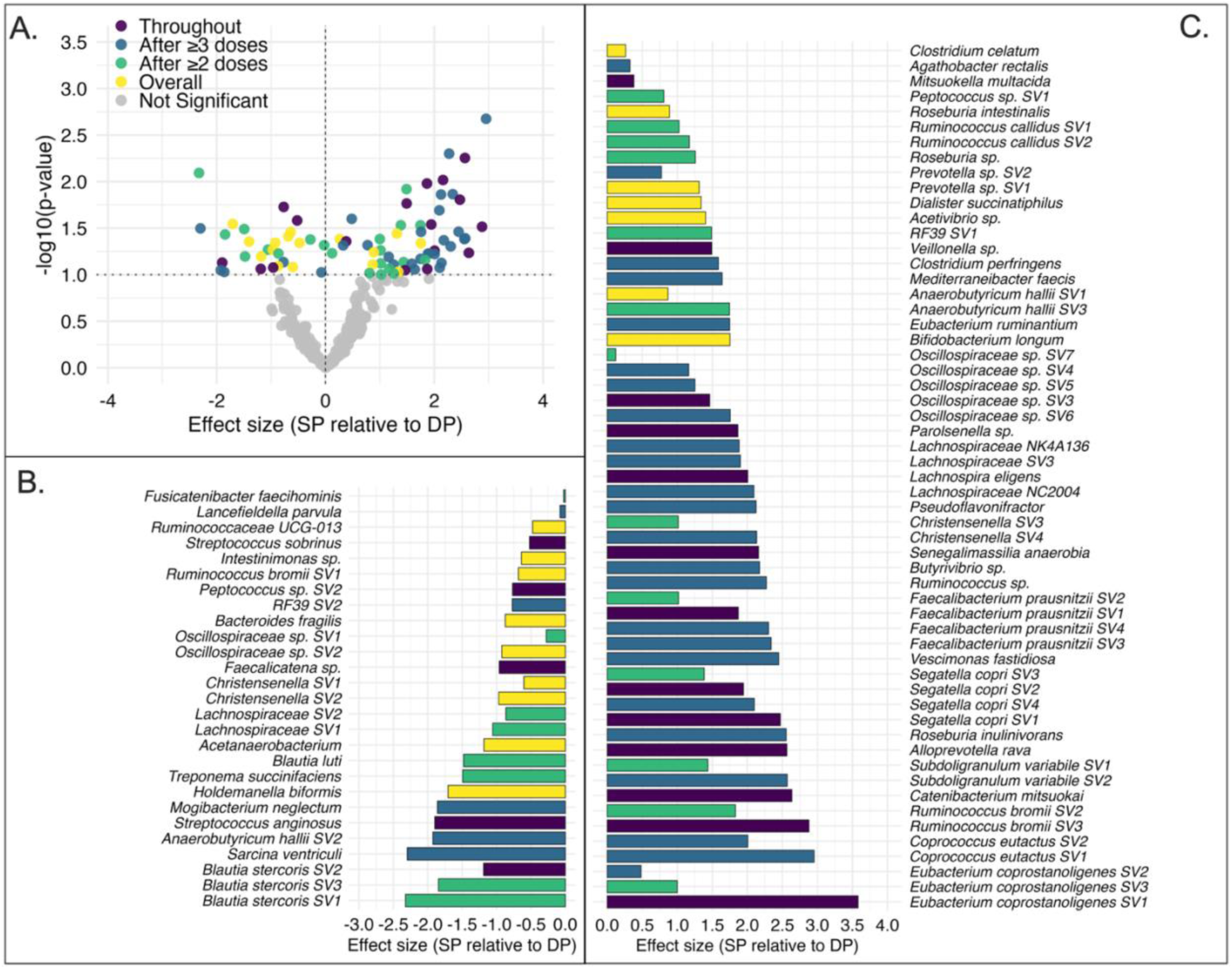
Differential abundance of gut microbiome sequence variants (SVs) by IPTp regimen (SP vs. DP). SVs passing sparsity filters were tested for differential abundance between SP and DP treatment arms using limma-voom and generalized linear mixed models (GLMM), applied across three analytical datasets: all IPTp samples, samples after ≥2 doses, and samples after ≥3 doses. **(A)** Volcano plot of effect size (log2 fold change, SP relative to DP) vs. −log10(p-value). Each point represents one SV. Colored points are significantly differentially abundant (n=84, p<0.1) and classified by whether they were significant across all three datasets (Throughout, purple); significant in samples after ≥3 doses (blue); significant in samples after ≥2 doses (green); significant in the pooled full follow-up dataset irrespective of dose (Overall, yellow); not significant (grey). **(B)** SVs reduced in SP relative to DP (n=27), ordered by taxonomy and effect size within taxon. **(C)** SVs enriched in SP relative to DP (n=57), ordered by taxonomy and effect size within taxon. Effect sizes are log2 fold changes from limma-voom or voom log2-CPM GLMM; for SVs significant only under ZINB-WaVE-normalized GLMM, the limma-voom effect size is shown with direction reflecting the significant GLMM result. Unadjusted p-values; no multiple testing correction applied.

Unadjusted p-values and a cut-off of p<0.1 were used given the exploratory, hypothesis-generating nature of the analyses. Support for the final set of differentially abundant SVs came from: i) concordance in the direction of relative abundance differences identified by the Gneiss geometric mean analysis and the formal differential abundance analysis for 77 of 84 significant SVs (91.7%); ii) downstream validation of biological plausibility driving these compositional changes, as described below (i.e., published reference genomes and metagenomes representing the taxa that changed with IPTp-SP vs DP clustered in a manner consistent with SP’s established microbial antifolate mode of action).

### Biological plausibility of SP-driven shifts in gut microbiota

SVs reduced with IPTp-SP (*vs*. DP treatment) included several pathobionts and opportunistic pathogens: *Streptococcus anginosus*, *S. sobrinus*, *Mogibacterium neglectum, Lancefieldella parvula*, and *Bacteroides fragilis* (Error! Reference source not found. **1B).** Among SVs enriched with SP treatment were colon commensals with reported short-chain fatty acid (SCFA)-producing and anti-inflammatory functions^18^: *Faecalibacterium prausnitzii*, *Segatella copri*, *Roseburia intestinalis*, *Mitsuokella multacida*, *Ruminococcus* sp., *Catenibacterium mitsuokai*, and *Butyrivibrio* sp (Error! Reference source not found. **1C)**. A few taxa with functions considered to be beneficial to host metabolism and gut barrier integrity (e.g., *Holdemanella biformis*, *Blautia luti*, *Ruminococcus bromii*)^18^ contributed SVs to both the reduced and enriched groups, underscoring strain-level complexity that 16S data cannot resolve. Not all SP-associated shifts were unambiguously beneficial: *Clostridium perfringens*, a toxin-producing enteric pathogen, was enriched with SP treatment, consistent with its ThyX-mediated antifolate tolerance (**Fig. 1C**, **Supplementary Note 2**). Additionally, seven taxa contributed SVs to both the enriched and reduced directions, suggesting strain-level heterogeneity that 16S resolution cannot distinguish (**Fig. 1B, 1C**). Thus, we performed functional analysis with publicly available strain-level reference and metagenome-assembled genomes (MAGs) from IMG^21^ corresponding to differentially abundant taxa to determine whether SP-associated microbial shifts were consistent with SP’s antifolate mode of action rather than stochastic drift.

Sulfadoxine and pyrimethamine competitively inhibit dihydropteroate synthase (DHPS) and dihydrofolate reductase (DHFR) respectively, two sequential enzymes in microbial folate synthesis, blocking production of tetrahydrofolate (THF). THF is the active folate cofactor required for nucleotide and amino acid biosynthesis. Bacteria that lack these targets, or that encode alternative routes to sustain these biosynthetic processes under antifolate pressure, are predicted to survive SP exposure (**Extended Data Fig. 2**). Of 53 differentially abundant taxa, 46 with genus- or species-level 16S classification were taken forward for genome retrieval, and prediction of SP susceptibility and plausible compensatory pathways for survival under SP pressure; seven taxa were excluded due to family-level or above resolution or contested polyphyletic genus assignment (**Extended Data Table 2).** High-quality isolate genomes representing each included taxon were annotated for the presence or absence of 15 genes relevant to bacterial folate metabolism, including the two SP targets DHPS and DHFR^22^ (**Methods**; **Extended Data Fig. 2**). Hierarchical clustering revealed 12 distinct folate functional clusters (**Fig. 2A**, **Extended Data Fig. 3**). Clusters differed by predicted SP susceptibility and survival strategy **(Fig. 2**, **Supplementary Note 2**).

**Fig. 2.**
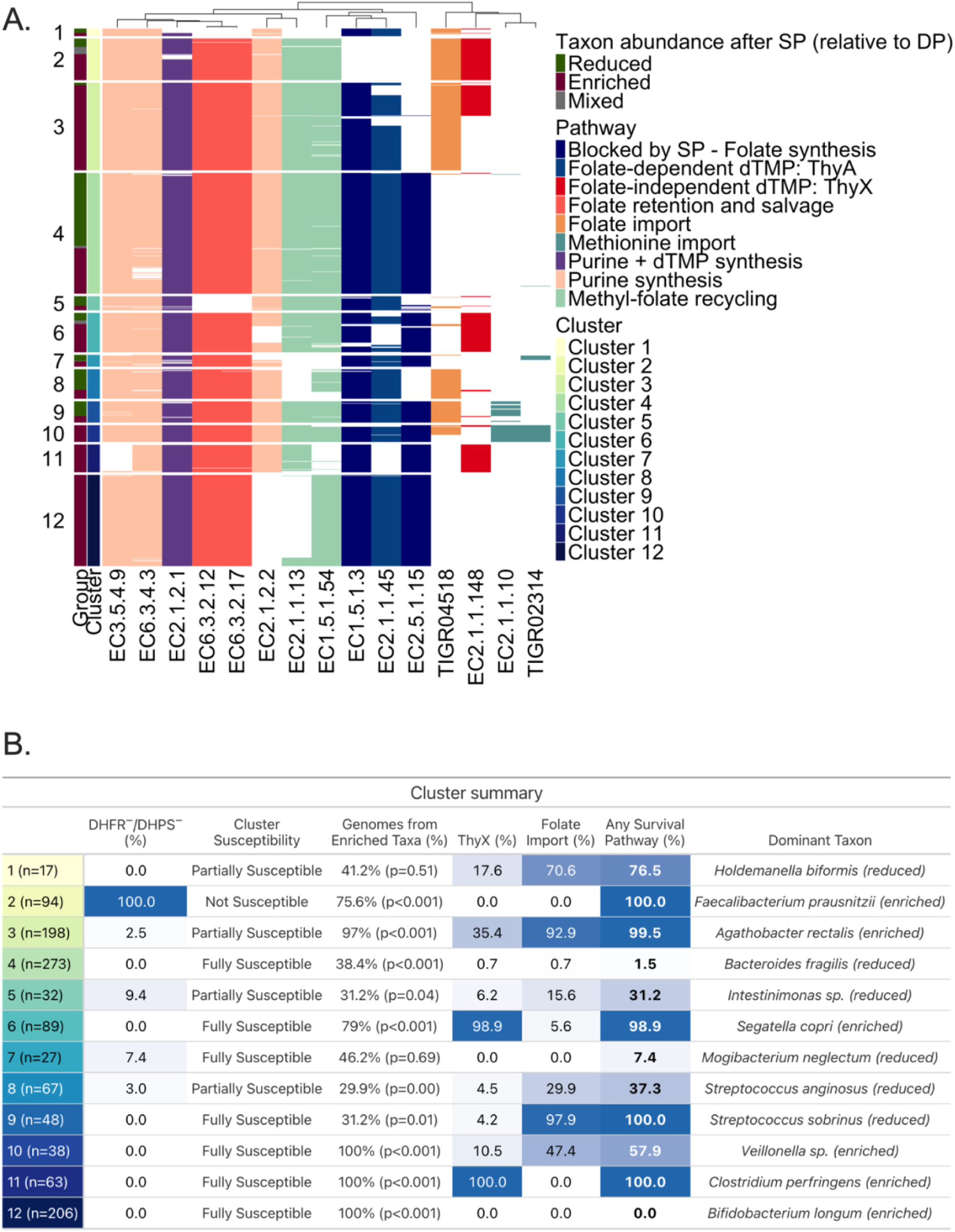
Gut microbiome taxa differentially abundant after IPTp-SP versus IPTp-DP cluster by folate pathway architecture, consistent with SP’s antifolate mode of action. **(A)** Heatmap showing the presence or absence of 15 folate and one-carbon metabolism gene functions, annotated by Enzyme Commission (EC) number or TIGRFAM accession, across 1,152 high-quality isolate genomes representing 46 differentially abundant taxa with species- or genus-level 16S classification. Genes are grouped by functional category: SP target enzymes (DHPS, EC2.5.1.15; DHFR, EC1.5.1.3); folate-independent dTMP synthesis (ThyX, EC2.1.1.148); folate-dependent dTMP synthesis (ThyA, EC2.1.1.45); folate retention and salvage (EC6.3.2.17, EC6.3.2.12); purine and dTMP biosynthesis (EC6.3.4.3, EC3.5.4.9, EC2.1.2.2, EC2.1.2.1); methyl-folate recycling (EC1.5.1.54, EC2.1.1.13); methionine import (EC2.1.1.10, TIGR02314); and folate import (TIGR04518). Genomes were hierarchically clustered (Ward’s method, Manhattan distance) based on binary gene presence/absence; k=12 was selected as the biologically interpretable solution following silhouette score evaluation across k=2-25 (Extended Data Fig. 3). Left-side annotations indicate whether each genome belongs to a taxon enriched (dark red), reduced (dark green), or mixed (grey) in SP- versus DP-treated women. **(B)** Cluster summary table showing per cluster: genome count (n); majority susceptibility category (not susceptible, partially susceptible, or fully susceptible based on DHPS and DHFR presence); percentage of genomes from enriched taxa with p-value from a within-cluster binomial test of the enriched:reduced split against a 50:50 null, Benjamini-Hochberg corrected across 12 clusters (Genomes from Enriched Taxa %); percentage of genomes carrying ThyX (ThyX %); percentage carrying a qualifying folate import pathway (Folate Import %); percentage carrying any survival mechanism (Any Survival Pathway %); and the dominant taxon with its observed RCT outcome. Clusters where a majority of genomes encode at least one survival mechanism tend to contain higher proportions of genomes from enriched taxa after IPTp-SP, consistent with antifolate selection pressure.

Predictions aligned with expected outcomes under SP’s established antifolate mode of action (**Fig. 2**). Taxa enriched after SP exposure (vs. DP) were disproportionately assigned to genome clusters lacking both SP targets (Cluster 1) or to genome clusters with one or both targets but with encoded folate import capacity (Clusters 3, 5) or folate-independent pathways for nucleotide synthesis via ThyX (Clusters 6, 7). Depleted taxa generally mapped to genome clusters that encoded DHPS and/or DHFR without compensatory mechanisms (e.g., Clusters 3, 4, 9, 10). The primary exception was Cluster 8, dominated by B. longum genomes, which lacks qualifying survival mechanism. *B. longum* persistence after SP vs. DP without a clear survival mechanism inferred from the 15 gene panel suggests other salvage dependencies not captured here or known cross-feeding with SP-favored taxa (*F. prausnitzii* and *S. copri*)^18^; this and other exceptions are discussed in **Supplementary Note 2**. At the taxon level, predicted survival capacity aligned with observed SP-enriched or SP-reduced outcome in 24 of 41 evaluable taxa (58.5%; taxa with a mixed observed outcome excluded; Supplementary Table 3). In a sensitivity analysis restricted to taxa with species-level 16S classification and more than three representative genomes, accuracy was 68.4% (13 of 19, **Extended Data Table 2**, **Supplementary Note 2**). These patterns support a mechanistic explanation consistent with SP’s antifolate selection pressure.

### SP-induced microbiome shifts mediate gestational weight gain

We previously demonstrated that SP’s beneficial effect on birthweight is mediated by increased gestational weight gain (GWG)^11^. This relationship has recently been replicated across trials by an independent meta-analysis^14^. We hypothesized that microbiome changes induced by SP partially mediate its effect on GWG. Consistent with our prior report of this cohort^11^, SP-treated women receiving ≥3 doses had 0.79 z-score units higher GWG than DP-treated women (β=0.79, CI95: 0.07-1.51, p=0.033, adjusted for baseline BMI; **Fig. 3A**). Among women with GWG measurements evaluated after ≥3 doses of IPTp (generally occurred within ∼1 month of delivery), 61 women (26 SP, 35 DP) were available for cross-sectional microbiome-GWG analyses after ≥2 doses, 49 (23 SP, 26 DP) after ≥3 doses, and 53 (22 SP, 31 DP) for longitudinal AUC-based analyses. A total of 16 SVs (15 unique taxa) associated most strongly with GWG in SP recipients (12 SP-enriched and 4 SP-reduced) (**Fig. 3B**, **Extended Data Fig. 4AB**). For 14 of 16 SVs, the strongest association with GWG was captured by cumulative AUC rather than cross-sectional analyses (**Extended Data Fig. 4B, Extended Data Table 3)**, consistent with the idea of sustained exposure (SP has a long half-life) and/or dose-dependent nature of SP’s microbiome effects.

**Fig. 3.**
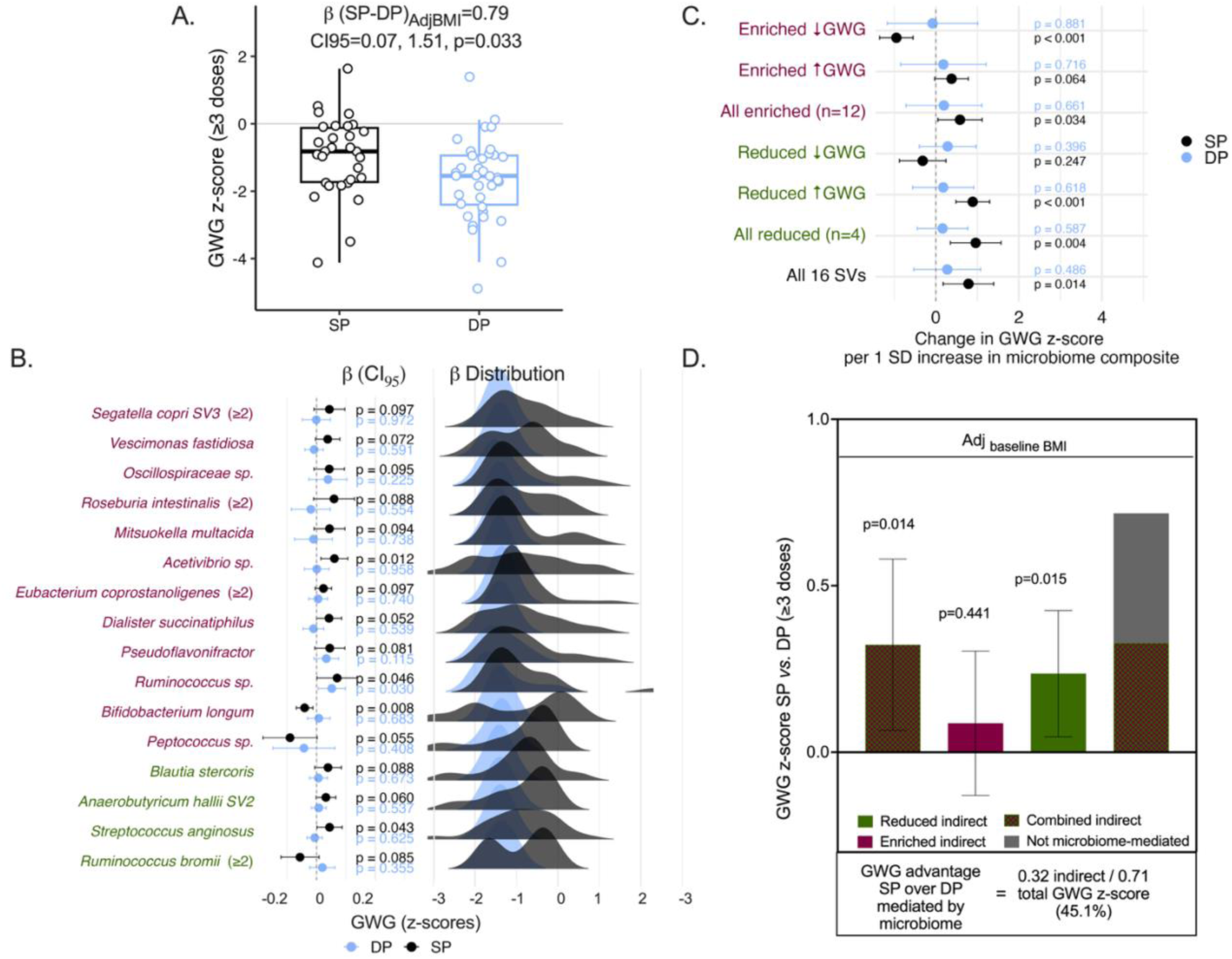
SP’s beneficial effect on gestational weight gain (GWG) is partly mediated by gut microbiome changes. **(A)** Gestational weight gain (GWG) z-scores by IPTp drug group among women receiving ≥3 doses with complete GWG and baseline BMI data (SP: n=30; DP: n=36). GWG z-scores were calculated using INTERGROWTH-21 standards. Scatter points show individual observed GWG z-scores; box plots show medians and interquartile ranges. The β coefficient was derived from a complete-case linear regression of GWG on drug group, adjusted for BMI at baseline (study entry). SP-treated women had significantly higher GWG than DP-treated women (β=0.79 GWG z-score units; 95% CI: 0.07-1.51; p=0.033). This replicates our previously published finding in this cohort in this microbiome-linked analytical subsample. **(B)** Associations between GWG and 16 SVs stratified by drug among women with ≥2 or ≥3 doses (SP: n=22-26; DP: n=31-35, varying by SV and analytical approach; see Extended Data Table 3). Univariable regressions (β ± 95% CI) and posterior density distributions of predicted GWG are shown for 16 of 84 differentially abundant SVs whose abundance trajectories correlated with GWG in SP but not DP recipients. Three SVs (*Anaerobutyricum hallii*, *Eubacterium coprostanoligenes*, and *Ruminococcus bromii*) were associated with GWG using cross-sectional relative abundance after ≥2 doses rather than AUC; all remaining SVs used cumulative area under the curve across visits. CLR-transformed relative abundances were sign-reversed prior to regression for reduced SVs in both drug groups (see Supplementary Note 2). SVs with p<0.1 were considered exploratory contributors to SP-specific GWG signal. SP-enriched taxa shown in maroon; SP-reduced in green. **(C)** Linear regression of GWG and composite microbiome scores formed by summing z-scored SVs within the indicated categories, among women with ≥3 doses (SP: n=23; DP: n=26). Separate regressions by drug group were run. Forest plots show the change in GWG z-score per 1-SD increase in each composite from SP- and DP-stratified models, with 95% CI and p-values (see Extended Data Table 4). **(D)** Causal mediation analysis estimating the proportion of SP’s total GWG advantage over DP attributable to SP-induced gut microbiome changes, adjusted for baseline BMI (SP: n=36; DP: n=41; larger than panel A because structural equation modeling with full-information maximum likelihood retains women contributing partial data on endogenous variables). The model was implemented in STATA19 using structural equation modeling. Bar heights show mediation effect estimates; error bars denote 95% CIs. Approximately 45% of SP’s total GWG advantage was mediated by SP-induced microbiome changes (combined indirect effect=0.32 GWG z-score units; 95% CI: 0.07-0.58; p=0.014), driven primarily by bacteria depleted in relative abundance with SP treatment (reduced indirect effect=0.24; 95% CI: 0.05-0.43; p=0.015); the enriched bacteria composite did not independently reach significance as a mediator (enriched indirect effect=0.09; 95% CI: −0.13-0.30; p=0.432). Full sensitivity analyses are reported in Extended Data Table 5.

Enriched taxa generally correlated with higher GWG, while reduced taxa correlated with lower GWG. Five exceptions were noted. Among enriched SVs, increases in *B. longum*, *Peptococcus sp*., and *Anaerobutyricum hallii* were associated with lower GWG. Among reduced SVs, higher abundance of *R. bromii* and *Lachnospiraceae* associated with higher GWG, suggesting that SP-driven depletion of these taxa may be detrimental rather than beneficial to GWG (**Fig. 3B**). We visualized participant-level predicted GWG as posterior density distributions and confirmed that the spread of individual predictions was consistent with the group-level patterns (**Fig. 3B**).

To test whether SP is associated with a microbiome configuration associated with higher GWG, we generated composite variables (as scores of centered log-ratio-transformed relative abundances) that combined the 16 SVs by whether they were reduced (n=4) or enriched after SP vs. DP (n=12), including whether they associated with lower or higher GWG, or all together (n=16). These groupings captured a combined microbiome signal in SP-treated women but not DP-treated women (**Fig. 3C**). The overall composite spanning all 16 SVs was positively associated with GWG in the SP group (p=0.014) but not in the DP group (p=0.486) (**Fig. 3C, Extended Data Table 4)**.

We used causal mediation analysis through structural equation modelling to quantify the extent of the GWG advantage over DP that could be attributed to SP-induced microbiome changes. Of SP’s (vs. DP’s) total advantage on GWG after ≥3 doses and adjusted for baseline BMI (mirror the GWG regression in Fig. 3A), ∼45% was mediated by the microbiome changes (indirect effect=0.32 GWG z-score units; CI95:0.07, 0.58; p=0.014, **Fig. 3D**, **Extended Data Table 5**). The indirect effect of 0.32 GWG z-score units corresponds to approximately 1.2 kg (95% CI: 0.3-2.2 kg), derived by multiplying the z-score estimate by the INTERGROWTH-21st standard deviation for gestational weight gain at 35-36 weeks’ gestation (σ=3.8 kg). This was driven primarily by the bacteria SP reduced relative to DP (indirect effect=0.24 GWG z-score; CI95:0.05, 0.43; p=0.015) rather than by the enriched bacteria (indirect effect=0.09 GWG z-score; CI95:-0.13, 0.30; p=0.432). The result was robust to individual and simultaneous adjustment for baseline BMI, gravidity, malaria infection, and socioeconomic status (**Extended Data Table 5**).

### SP → microbiome-attributable GWG → birthweight segment

Our secondary hypothesis was that microbiome-mediated GWG associates with birthweight (grams). Observed GWG was positively associated with birthweight in SP-treated but not DP-treated women **(Fig. 4A**), consistent with our prior report of this cohort^11^. Next, we tested how the effect of the microbiome-mediated GWG related to birthweight in SP and DP recipients. Analyses included 70 mother-infant dyads with linked GWG, birthweight, and maternal microbiome data after ≥3 IPTp doses (SP: n=34, DP: n=36). For each participant, microbiome-predicted GWG was derived from multivariable regression of observed GWG on the four benefit-coded composite scores within each drug arm (**Fig. 3C);** these predicted values were then evaluated for their association with birthweight, separately within each arm.

**Fig. 4.**
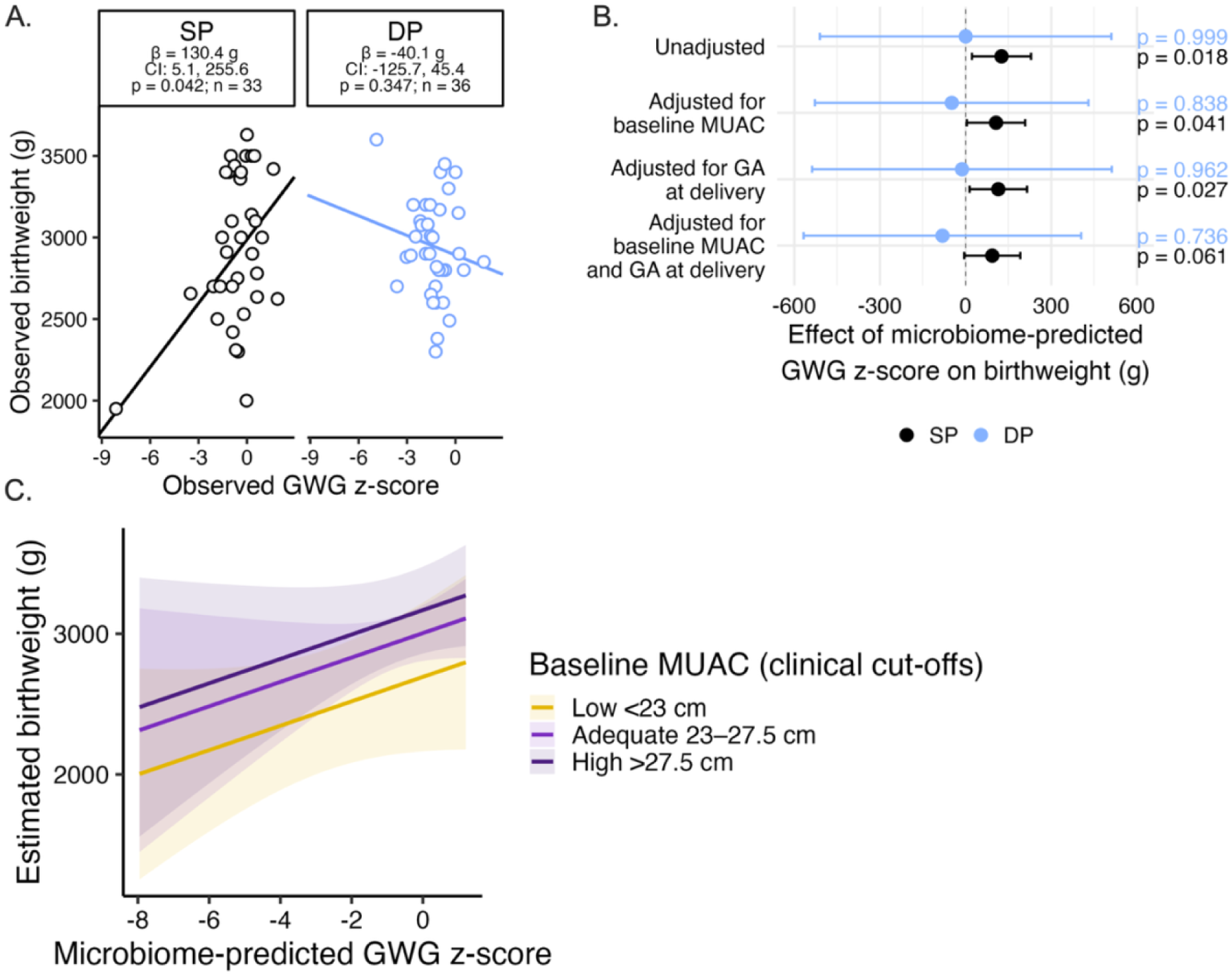
Microbiome changes that causally mediated SP’s gestational weight gain (GWG) benefit associate with higher birthweight. **(A)** Relationship between observed GWG z-score and observed birthweight (g) among women receiving ≥3 IPTp doses (SP: n=34, DP: n=36). Points show individuals; lines show drug-stratified linear regression fits derived from STATA; inset reports the slope (g birthweight per 1 SD of GWG z-score) and p-value for each group. **(B)** The microbiome changes shown to causally mediate SP’s GWG advantage were associated with higher infant birthweight in SP but not DP recipients. Sixteen SVs were grouped into four benefit-coded composite predictors based on SP-induced abundance changes and their GWG associations (see **Supplementary Note 2**); multivariable regression of observed GWG on these composites within each drug arm generated participant-level microbiome-predicted GWG values (Fig. 3C), which were then tested for association with birthweight. In SP-treated women, higher microbiome-predicted GWG was significantly associated with increased birthweight after ≥3 doses (+126.0g; 95% CI: +22.6, +229.3; p=0.019; SP: n=34, DP: n=36). Effects remained stable after adjustment for gestational age at delivery and baseline nutritional status by MUAC, but confidence intervals widening under full adjustment for both MUAC and gestational age at delivery (**Extended Data Table 6**). These effects were not observed in DP-treated women. **(C)** Maternal MUAC at enrollment stratified but did not modify the relationship between microbiome-predicted GWG z-score and birthweight in SP recipients (interaction p=0.992). Regression lines (shaded 95% CIs) show estimated birthweight across microbiome-predicted GWG z-score in SP recipients, stratified by clinically defined MUAC cut-offs (low <23 cm; adequate 23-27.5 cm; high >27.5 cm). The consistent slopes across MUAC strata indicate that the microbiome-GWG-birthweight association is independent of baseline maternal nutritional status.

In SP recipients, a 1 SD increase in microbiome-predicted GWG z-score was significantly associated with higher birthweight after ≥3 doses, the number of doses recommended by the WHO (+126.0g; 95% CI: +22.6, +229.3; p=0.019; **Fig. 4B**; **Extended Data Table 6**). Effects remained stable after adjustment for gestational age at delivery and baseline nutritional status by MUAC across both dose groups (**Extended Data Table 6**). The birthweight point estimate was stable after adjustment for gestational age at delivery and baseline nutritional status by MUAC, though confidence intervals widened under simultaneous adjustment for baseline MUAC and gestational age at delivery (β=93.5g; 95% CI: −4.7, 191.7; p=0.061; **Extended Data Table 6**), reflecting the limited power of this exploratory sample for fully adjusted analyses

No corresponding associations were observed in DP recipients at either dose threshold (**Extended Data Table 6).** Baseline MUAC independently stratified birthweight outcomes (**Fig. 4C**). MUAC was not associated with microbiome-predicted GWG in either arm, nor did it modify the microbiome-predicted GWG-birthweight relationship (interaction p=0.992). Mothers with higher MUAC at enrollment delivered heavier infants across both treatment groups (**Fig. 4C).** Together, these findings support two parallel contributors to birthweight in this cohort: an SP-linked microbiome signal that tracks with GWG and birthweight, and baseline maternal nutritional status that shifts birthweight upward regardless of treatment arm.

## Discussion

Despite widespread drug resistance that has compromised SP’s antimalarial efficacy, IPTp-SP continues to deliver consistent and reproducible improvements in birthweight outcomes, outperforming more potent non-antibiotic antimalarials like DP for fetal growth outcomes^5, 6^. This paradox points to a malaria-independent mechanism^23^, which remains unexplained. Leveraging a randomized controlled trial comparing IPTp-SP to IPTp-DP in pregnant women from Malawi that enabled causal inference, we previously identified that gestational weight gain (GWG) mediates approximately two thirds of SP’s positive effect on birthweight^11^. A subsequent individual participant data meta-analysis of IPTp trials corroborated the effect of SP on GWG^14^. We now present the first evidence that a malaria-independent biological pathway through which SP improves GWG and birthweight is through the maternal gut. Compared to the non-antibiotic comparator arm, IPTp-SP changed the composition of the maternal gut microbiome during the 2^nd^ and 3^rd^ trimesters when GWG and fetal growth accelerate and peak in a manner consistent with its known pharmacology. Microbiome reconfiguration by SP causally mediated 45% of SP’s total GWG advantage over DP (β=+0.32, 95%CI 0.07–0.58, p=0.014), adjusted for baseline BMI and stable across pre-specified sensitivity analyses incorporating gravidity, malaria, socioeconomic status, and additional covariates (range 44–51%). This corresponded to approximately 1.2 kg of the 2.4 kg additional GWG observed in SP versus DP recipients and coincided with 93-126g higher birthweight in babies born to SP but not DP recipients, independent of gestational age and maternal nutritional status.

The compositional changes to the maternal gut bacterial microbiome were consistent with SP pharmacology over its long half-life (∼200h for sulfadoxine, ∼100h for pyrimethamine^17^) and established antifolate molecular mode of action^8^. SP is readily absorbed in the small intestine with high but incomplete bioavailability (>90% in healthy adults)^16, 17^. The unabsorbed fraction, theoretically ∼10% of the administered dose, is estimated to generate luminal concentrations in the range of reported antibacterial minimal inhibitory concentrations (∼4-24 µg/mL)^24^ before elimination in feces^16^. The unabsorbed portion may be higher in pregnancy, which alters SP pharmacokinetics^17^. SP’s elimination is primarily renal^16^, though sulfadoxine has a minor biliary excretion component that delivers it back to the small intestine from the liver through enterohepatic circulation^16^, providing an additional route of intestinal drug exposure over the long half-life.

We identified 53 unique taxa (84 sequence variants, SVs) that differed in relative abundance among SP vs. DP microbiomes: 15 were exclusively reduced, 31 taxa were exclusively enriched with SP vs. DP, and 7 taxa contributed SVs to both enriched and reduced groups. Using strain-level functional genomics across 1,152 high-quality isolate genomes representing reduced and enriched taxa, we found the on-treatment shifts in microbiome composition to be consistent with SP’s antifolate antibiotic activity rather than stochastic drift. SP inhibits two key steps in microbial folate synthesis catalyzed by the bacterial enzymes DHPS and DHFR^8^. Folate is indispensable to the survival of all living things, serving as an essential cofactor in the synthesis of nucleotides required for DNA replication and cell division^8^. Bacteria that rely on *de novo* folate synthesis are susceptible to SP and, in the absence of compensatory mechanisms (e.g. alternative folate acquisition pathways and/or metabolic pathways that bypass folate for nucleotides), are expected to be suppressed. We observed that published strains representing taxa depleted under SP (vs. DP) encoded DHPS and/or DHFR and lacked compensatory mechanisms (e.g. *Bacteroides fragilis*, *Streptococcus anginosus*, *S. sobrinus*). Strains of taxa that expanded under SP either lacked *dhps* and *dhfr* altogether (e.g., *Faecalibacterium prausnitzii*) or encoded alternative survival strategies that bypass antifolate pressure, of which ThyX was the most notable example (e.g., *Segatella copri)*.

Among reduced taxa, a set are recognized gastrointestinal pathobionts. *B. fragilis* comprises toxigenic strains causing colitis, inflammatory diarrhea, and systemic inflammation ^18, 25–27^. *S. anginosus*, *S. sobrinus*, *Mogibacterium neglectum*, and *Lancefieldella parvula* are oral-origin pathobionts capable of ectopic small bowel colonization^28–32^. The small bowel is where 90% of all host-digestible calories are absorbed and typically has a low microbial biomass^33^. Chronic small intestinal colonization with oral pathobionts is a recognized microbial feature of LMIC children with growth shortfalls and enteric dysfunction (ED)^28–32^. ED enteropathy^30, 34^ mirrors that of small intestinal bacterial overgrowth (SIBO) in high income countries^33^. In SIBO, excessive bacteria drive mucosal inflammation that damages absorptive villi or directly interfere with host nutrient absorption through specialized virulence factors^33^. The same inflammatory and impaired nutrient handling pathways are now being investigated at a mechanistic level as causal drivers of childhood ED and stunting ^28–32^. Clinically-established consequences of SIBO include fat malabsorption syndromes and deficiencies in fat-soluble vitamins and B12^35^, independent risk factors for adverse fetal growth outcomes, including LMIC pregnancies^36, 37^. Whether enteropathies analogous to infant ED characterize LMIC pregnancies complicated by inadequate GWG or adverse fetal growth outcomes is under-explored^34^. Our previous report of high maternal carriage rates of enteropathogens commonly detected in stunted children with ED^38^, which tracked with lower GWG and birthweight^11^, supports this idea. Studies of paired stool-duodenal aspirates show that small intestinal bacteria characteristic of this ED pathology can be detected in stool^28, 31^. SP-mediated suppression of this pathobiont community may therefore contribute to alleviation of enteropathy, inflammation, and nutrient malabsorption, pathways plausibly linking the maternal gut microbiome to GWG and fetal growth outcomes.

SP also favored reciprocal expansions in anaerobic colonic commensals, whose known biology complements the possible anti-inflammatory and metabolic benefit of pathobiont suppression. Among the 31 enriched taxa, 74% (23/31) are recognized primary degraders of dietary fiber or resistant starch (e.g., *Ruminococcus*) or producers of short-chain fatty acids butyrate, acetate or propionate (e.g., *Faecalibacterium*, *Roseburia*, *Agathobacter*, *Subdoligranulum, Segatella, Prevotella*, *Mitsuokella*, *Dialister, Bifidobacterium*)^18^. SCFA producers are enriched in optimal colon communities^18^. SCFAs are microbial metabolites and signaling molecules with pleiotropic functions that link the gut to host metabolism and immunity^18^. They can be utilized directly by host tissues as energy substrates^39^, and approximately 5-10% of total daily caloric requirement and metabolizable energy in humans can be met by these SCFAs when produced in the colon through fermentation of dietary fiber^39^. Because Malawian diets are predominantly plant-based and rich in complex carbohydrates^40^, SP-associated enrichment of carbohydrate-fermenting taxa is biologically coherent in this dietary context. Through inhibition of histone deacetylases and binding of G-protein-coupled high-affinity host receptors, SCFA exert anti-inflammatory effects far beyond the gastrointestinal tract^18^. Depletion of butyrate-producing bacteria is a consistent cross-country fecal signature of ED in stunted LMIC children, alongside nutrient malabsorption^32^. Because mechanistically butyrate producers depend on acetate producers as their substrate source^18^, SP increasing both guilds simultaneously suggests functional gut changes beyond taxonomic rearrangements. These enrichments are a notable exception to antibiotic-associated dysbiosis, consistent with microbiome-sparing effects of co-trimoxazole, another antifolate antibiotic in clinical use^41^. However, not all SP-associated enrichments were unambiguously beneficial. Notably *C. perfringens*, a toxin-producing enteric pathogen, was enriched, possibly via ThyX-mediated survival under antifolate pressure. To our knowledge, this is the first dedicated characterization of any antifolate antibiotic on the human gut microbiome in pregnancy.

Sixteen (16) differentially abundant bacteria (12 enriched and 4 depleted) associated most strongly with GWG among SP but not DP recipients (∼30%, 16/53). Their combined effects explained almost half of SP’s total GWG advantage over DP (∼1.2 kg of the 2.4kg total by 36 weeks’ gestation), which in turn predicted babies born to SP mothers to be 93-126g heavier that babies of DP mothers, irrespective of mothers’ malaria and nutritional status at baseline or baby’s gestational age at delivery. Our observed birthweight effect sizes match the overall 71g-161g benefit reported in the most recent meta-analysis of IPTp-SP efficacy across sub-Saharan Africa^6^. These results identify a subset of bacteria that in combination are net favorable for GWG and birthweight, even though not every individual taxon shifted in a clearly beneficial direction. This functional signature is also seen in an independent setting characterized by undernutrition and enteric pathogens, albeit infants with ED rather than pregnant women. In malnourished Bangladeshi infants at risk of growth shortfalls but successfully treated with the first microbiota-directed therapeutic food shown to outperform higher-calorie standard interventions^19, 29^, taxa that predicted poorer growth in these children^19, 29^ also predicted lower GWG in our cohort *(S. anginosus*, *B. longum*); conversely, taxa positively correlated with ponderal growth in these children and the bacterial targets engaged by the intervention to drive recovery from malnutrition^19, 29^ were positive predictors of GWG in our cohort (*S. copri*, *R. intestinalis*, *M. multacida*). Resistant starch-degrading taxa were also the predominant predictors of infant birthweight in an independent longitudinal cohort of rural Zimbabwean mothers^42^, extending this functional signal to a second pregnant LMIC population.

The 16 taxa in our signature could be interpreted as a metabolic and ecological endotype characterized by a gut community that minimizes intestinal inflammation and improves nutrient availability. GWG is an integrated phenotype of maternal metabolic adaptation, encompassing maternal fat accretion, plasma volume expansion, placental growth, and fetal growth, each driven by maternal metabolic adaptations that are themselves sensitive to gut inflammatory status and nutrient availability^43^. Reducing *S. anginosus* plausibly alleviates small intestinal pathobiont-driven inflammation and nutrient malabsorption. The clinical efficacy of this endotype may further involve a balanced acetate-butyrate network, in which keystone primary fiber degraders (e.g., *S. copri*) produce acetate required by secondary consumers (e.g., *Roseburia intestinalis, Mitsuokella multacida,* and *Pseudoflavonifractor sp*.) to produce butyrate aromatic amino acids, or B-vitamins, energy substrates usable by the host in anabolism^18^. *Anaerobutyricum hallii*, while butyrogenic, has a unique requirement for excess acetate that under conditions of dietary scarcity can deplete the acetate pool^44^. Its reduction by SP is consistent with preservation of acetate availability for the host, which has been positively linked to GWG^45^.

Our microbiome-mediated pathway accounts for approximately 45% of SP’s GWG advantage, leaving the remainder unexplained. Recent data from a human intestinal chip model of nutritionally induced enteric dysfunction demonstrated that SP directly alleviates mucosal inflammation independent of microbial activity^46^, raising the possibility that SP exerts parallel direct anti-inflammatory effects on the gut epithelium. This is consistent with pyrimethamine’s direct immunomodulatory effect on the human host, including human STAT3 signaling^47^ and low-affinity activity on human DHFR^48^. This could be a complementary pathway that may account for part of the remaining GWG benefit and potentially for the portion of SP’s birthweight effect that does not operate through GWG.

Seeing overlaps in modifiable “weight-promoting” microbiome patterns between pregnant Malawian women and malnourished Bangladeshi children treated with a dietary intervention raises the question of whether IPTp-SP is addressing an intestinal pathway of impaired growth potential shared across populations and life stages burdened by undernutrition and enteropathy, and whether SP’s gains could be amplified with nutritional co-interventions. Our result that maternal nutritional status (MUAC) at enrollment stratified birthweight outcomes in both drug groups supports this idea.

Our study has limitations. The sample size, while consistent with longitudinal pregnancy microbiome studies employing repeated-measures designs, limits statistical power for subgroup analyses and formal moderated mediation modeling. Taxonomic resolution was limited to 16S amplicon SVs, and strain-level heterogeneity likely underlies many of the observed effects, warranting future strain-resolved analyses. Potential detrimental effects of antimicrobial exposure on the early-life microbiome, including depletion of beneficial commensals, pathobiont enrichment, and selection of resistance determinants, also require dedicated study. In prior work in this cohort, two or more doses of IPTp-SP were associated with a selective increase in the antifolate resistance gene *dfrA17*, but not with broad changes in the maternal gut resistome^49^. Because the cohort was restricted to Malawian women, generalizability remains to be tested. All findings should be considered exploratory and hypothesis-generating pending replication in larger, independent cohorts.

These findings position maternal intestinal enteropathy and microbial metabolism as modifiable targets in the pathway from impaired maternal gut function to reduced GWG, and fetal growth. Future studies are needed to determine whether these microbial changes translate to measurable shifts in maternal metabolic capacity and placental nutrient transfer that explain improved fetal growth outcomes.

## Methods

### Ethics Statement

This study was reviewed and approved by the Institutional Review Boards at the University of North Carolina at Chapel Hill (#16-1260), the Centers for Disease Control and Prevention (#6836), and the College of Medicine Research and Ethics Committee (COMREC), University of Malawi (#P.02/16/1872). All participants provided written informed consent. The trial is registered at ClinicalTrials.gov (NCT03009526).

### Study population

The present study is an ancillary investigation to an open-label, 2-arm randomized controlled superiority trial (RCT) designed to compare the efficacy and safety of monthly administration of either sulfadoxine-pyrimethamine (SP) or dihydroartemisinin-piperaquine (DP) for intermittent preventive treatment of malaria in pregnancy (IPTp) in Machinga district, Malawi (ClinicalTrials.gov Identifier: NCT03009526). Pregnant women between 16- and 28-weeks gestational age attending their first antenatal care (ANC) visit at Machinga District Hospital were recruited. Criteria for inclusion were informed consent for both parent study and sub-study, viable pregnancy, no history of IPTp use in the current pregnancy, a negative HIV test, and residency in the study area. Women with high-risk pregnancies or other medical conditions were excluded. Gut microbiome changes following receipt of DP or SP were a prespecified secondary objective of the parent randomized trial NCT03009526. The specific differential abundance, mediation, and microbiome-predicted birthweight analyses presented here were not separately prespecified in the trial registry and are reported as exploratory analyses within that broader microbiome objective.

### Gestational weight gain measurements and definitions

At each visit, including enrollment, women were weighed on a digital scale, accurate to the nearest 0.1 kg. Gestational weight gain (GWG) was estimated as previously described using the INTERGROWTH-21st Project standards ^11^. In brief, GWG was first expressed as the change in kg relative to baseline weight at study enrollment. To account for pre-pregnancy weight, we added the average expected INTERGROWTH-21st GWG (based on gestational age at enrollment) to each woman’s weight gain prior to z-score calculation using the INTERGROWTH calculator (https://intergrowth21.tghn.org/gestational-weight-gain/~c6). GWG z-score was used as a continuous variable.

### Fecal sample collection and pathogen detection

Fecal specimen collection, processing, shipping, and DNA isolation were previously described ^11^. In brief, at enrollment, which was the first antenatal visit for the current pregnancy, mothers were randomized to either the SP or DP arm and provided a baseline fecal specimen. Thereafter, they were scheduled for monthly antenatal study visits, each 28-30 days apart. At each study visit, fecal specimens were collected and IPTp was administered. Some women did not bring stool samples at each visit, but all women received the drug as scheduled. Upon submission to the study nurse, stool samples were taken to the field laboratory in Machinga, partitioned by laboratory staff into 5 equal aliquots in a biological cabinet using ethanol-treated surfaces and spatulas, and placed in sterile 2mL polypropylene cryotubes. The specimen aliquots were then placed in a −80°C freezer for storage. Three of the 5 aliquots were shipped on dry ice to UNC at Chapel Hill, United States for further laboratory procedures, including bacterial gut microbiome profiling by 16S rRNA gene amplicon sequencing targeting the V4 hypervariable region. Upon arrival at UNC at Chapel Hill, frozen stool samples were placed in −80°C freezers until DNA extraction. DNA was extracted from thawed stool samples in batches of 23 with one negative control (PBS).

### Amplification of 16S ribosomal RNA (rRNA) V4 variable region

Each fecal DNA sample was amplified at the V4 variable region of the 16S rRNA gene in two independent PCR replicates. Each PCR replicate was sequenced. Each sequencing batch per flow cell included two positive controls (ZymoBIOMICS Microbial Community DNA Standard) and two negative controls (one negative control represented one randomly selected DNA extraction negative control from 24 available and one PCR negative control which received all PCR components except for gDNA).

For PCR, we used the forward primer 515, 5’-GA GTG CCA GCM GCC GCG GTA A-3’, and reverse primer 806, 5’-ACG GAC TAC HVG GGT WTC TAA T-3’. Both the forward and reverse primer incorporated frameshifting nucleotides of six different lengths each; frameshifting has been demonstrated to enhance library diversity and thus improve the depth and quality of sequence data. Each of the 16S rRNA PCR products were quantified using a Qubit 3.0 fluorometer. Up to 192 amplicons were pooled equimolarly per flow cell for sequencing. In the case of negative controls with undetectable signal of double-stranded DNA by Qubit fluorometer, we normalized the input volume to the lowest concentration amplicon derived from a fecal gDNA sample. The final pooled sequencing library was quantified by quantitative real-time PCR (qPCR) using the Kapa Library Quantification and the distribution of fragment lengths confirmed by TapeStation using the High Sensitivity D1000 kit (Agilent). Sequencing was performed using 2×250 Illumina MiSeq flow cells at the High-Throughput Sequencing Facility at the UNC School of Medicine.

### Sequencing reads processing

Sequence processing and analysis was conducted in QIIME2 version 2019.7^50^. First, raw paired-end fastq reads from each sequencing run were demultiplexed and quality-filtered with the q2-demux plugin, followed by removal of sequencing adapters and PCR primers. Next, sequences were denoised with dada2 ^51^ independently for each sequencing run before merging into a full dataset. SVs were aligned with mafft^52^ (via q2-alignment).

### Validation of sequencing controls and sample replicate concordance

To quantify sources of PCR or sequencing bias or contamination, we first verified the composition of sequences identified in negative control samples. Next, we compared taxa frequencies of sequences originating from the mock bacterial community positive control (ZymoBIOMICS Microbial Community Standard) against the expected, theoretical composition. Finally, because duplicate 16S amplicons were generated from each gDNA sample and sequenced independently, we investigated differences between sample replicates by assessing differences in α-diversity metrics (number of observed taxa, Shannon’s diversity index, and Faith’s Phylogenetic Diversity) and β-diversity metrics (Bray-Curtis distance, unweighted UniFrac distance, and weighted UniFrac distance), as a function of drug group. Differences in α-diversity between replicates were assessed using q2-longitudinal and Wilcoxon rank sum tests and their distribution visualized by scatterplots.

Mann-Whitney U tests were used to assess whether paired differences were significant between drug groups. Differences in β-diversity between replicates, as a function of drug group, were evaluated using q2-longitudinal and Mann-Whitney U tests.

### Defining amplicon sequence variants (SVs) from sample replicates

We leveraged having sequenced two amplicon replicates per sample to define final amplicon sequence variants (SVs). For each sample, we performed an intersection of observed features, and we retained only the ones observed in both replicates. Final SVs were then excluded from downstream analyses if: 1) they occurred at a threshold lower than 0.1% of all SVs in the dataset; 2) they were observed in fewer than 5 samples; and/or 3) they had a read count less than 50. The remaining SVs were assigned a taxonomy using the q2-feature-classifier and the 138 release of the SILVA reference database at 99% sequence similarity. Phylogeny was done with fasttree2 ^53^ (via q2-phylogeny). Sequences mapping to phylum Cyanobacteria, family of mitochondria, and class of chloroplast were removed. Metrics of ɑ- and β-diversity were estimated using q2-diversity after samples were rarefied (subsampled without replacement) to 51,000 reads/sample, as appropriate.

### Global Microbiome Diversity Analyses

We assessed whether IPTp altered overall gut microbial diversity as measured by 16S rRNA gene amplicon sequencing. Alpha-diversity was evaluated using Shannon’s index and beta-diversity was calculated using weighted UniFrac distances and visualized with principal coordinates analysis (PCoA). We conducted two complementary analyses. First, for longitudinal within-group comparisons, we used linear mixed-effects models (qiime longitudinal linear-mixed-effects) to evaluate changes in Shannon diversity over time by treatment group. For beta diversity, we assessed changes from baseline using two-sided Mantel tests and pairwise PERMANOVA (999 permutations). Second, for cross-sectional comparisons, we assessed alpha and beta diversity between drug groups at each follow-up visit independently to test for dose-dependent effects of IPTp. Differences in beta diversity were evaluated using PERMANOVA.

### Compositional analysis exploration using Gneiss

To explore treatment-associated differences in bacterial relative abundance, we first applied the Gneiss tool in QIIME 2 to centered log-ratio (CLR) transformed SV abundance data. This method applies correlation-based hierarchical clustering to group SVs into balances of co-varying taxa (taxa that go up and/or down together in characteristic ways) for each drug group. Within each balance, SVs discriminating most strongly between SP and DP microbiomes were identified: for each SV, we computed the log_e_ difference in geometric mean abundance between treatment groups to rank them by effect size and identify the ones that contribute the most to the balance’s ability to SP and DP microbiomes apart.

### Formal differential abundance testing

#### Data preparation for differential abundance testing

Sparse SVs with zero abundance in ≥80% of samples in both drug groups were excluded, as sparsity impact reliability for differential abundance or regression analyses with clinical variables. Sparsity was evaluated separately by drug group to avoid discarding SVs genuinely enriched or depleted in one arm relative to the other, a central hypothesis of this study.

ZINB-WaVE (zero-inflated negative binomial) was applied to raw SV counts with K=0 and observational weights enabled, as recommended for weight generation when group membership is known *a priori*^54^. ZINB-WaVE models the zero-generating process explicitly, assigning each observation a weight reflecting the probability that a zero represents true biological absence rather than a technical dropout. The resulting observational weights were incorporated into the limma-voom pipeline to stabilize variance estimates. For limma-voom, these weights were incorporated during voom transformation to stabilize variance estimates. For the GLMM, ZINB-WaVE normalized values were used as the response variable with a Gaussian error distribution, handling zero-inflation upstream of the model. After ZINB-WaVE (zero-inflated negative binomial) was applied to raw SV counts with K=0 and observational weights, SVs differentially abundant at the pre-treatment baseline visit were identified using limma-voom (p<0.05, one observation per participant, no random effects),via the edgeR and limma packages^55^). These were excluded prior to main differential abundance testing.

*Differential abundance testing by IPTp group (SP vs. DP)*

To identify SVs differentially abundant by IPTp group (SP *vs*. DP), we applied two independent methods, limma-voom with TMMwsp and generalized linear mixed models (GLMM, using the glmmTMB R package). For limma-voom, SV counts were normalized using the trimmed mean of M-values with singleton pairing (TMMwsp) method and voom-transformed with ZINB-WaVE weights incorporated^56^. GLMMs and linear regression models were fit using both voom-normalized log2-CPM values and ZINB-WaVE normalized values. Voom log2-CPM values place GLMM coefficients on the same scale as limma-voom log2 fold changes, enabling direct comparison of effect sizes across methods; ZINB-WaVE normalized values follow published recommendations for GLMM-based differential abundance testing in microbiome data^54^. Three analytical datasets were assessed: all IPTp-period samples (repeated measures); all samples per participant collected after ≥2 doses (repeated measures); and final samples collected after ≥3 doses (one per participant, cross-sectional). For limma-voom, repeated measures were addressed using the duplicateCorrelation() function with participant ID as a blocking factor; a weighted linear model was then fit using lmFit(), followed by empirical Bayes moderation with eBayes().limma-voom was the principal method based on strong benchmarking performance^56^. GLMM served as an independent check. Each participant contributed 1-4 samples in longitudinal analyses; no analysis treats repeated observations as independent.

An SV was considered differentially abundant if: (1) p<0.1 by at least one method in at least one analytical dataset; and (2) effect direction was consistent across all statistically significant results. SVs with conflicting directions in effect size were excluded. Unadjusted p-values (p<0.1) were used given the exploratory nature of these analyses.

SVs were classified by the analytical dataset in which significance was detected. SVs significant only in the pooled full follow-up dataset (irrespective of dose) were labeled *Overall*; those significant after ≥2 doses were labeled *After ≥2 doses*; those significant after ≥3 doses were labeled *After ≥3 doses*; and those significant across all three datasets were labeled *Throughout*. SVs in the dose-stratified categories suggest effects that emerge or strengthen with cumulative dose exposure.

Log2 fold changes from limma-voom were used for visualization; where only GLMM was significant, the GLMM effect size was shown. Final results were visualized in volcano plots.

### Folate Gene Mining and Functional Annotation

Sulfadoxine competitively inhibits microbial dihydropteroate synthase (DHPS), and pyrimethamine competitively inhibits microbial dihydrofolate reductase (DHFR)^8^. DHPS and DHFR catalyze two sequential and essential steps in the de novo synthesis of tetrahydrofolate (THF), the active folate required for DNA, purine, and methionine biosynthesis (**Extended Data Fig. 2).** Microbes that rely on synthesizing THF *de novo* are therefore vulnerable to disruption of these steps unless they can obtain folate or folate derivatives through import, salvage, or retention pathways. We hypothesized that taxon-specific differential abundance patterns in SP-exposed mothers reflect this antifolate mode of action. Because 16S amplicon data lack strain-level resolution and no new genomes were generated from our cohort, we retrieved publicly available isolate genomes from the Integrated Microbial Genomes (IMG) database and additional public sequence repositories. Genomes were filtered to retain high-quality assemblies (IMG High Quality flag=Yes; Sequencing Status: Finished or Permanent Draft); metagenome-assembled genomes and unscreened single-cell amplified genomes were excluded.

Only differentially abundant taxa resolved to the genus or species level were included; taxa with family-level or above 16S assignments, or belonging to contested polyphyletic genera, were excluded to avoid misattributing gene content. Taxa with SVs detected in both enriched and reduced directions (mixed designation) were retained in the analysis but excluded from prediction accuracy evaluation, as no unidirectional ground-truth outcome exists for comparison. Guided by established biological roles in folate and one-carbon metabolism (KEGG map00790/map00670028), we curated an *a priori* panel of 15 gene functions expected to shape a microbe’s ability to cope with THF disruption, including DHPS (EC2.5.1.15) and DHFR (EC1.5.1.3): ThyX (EC2.1.1.148); folate import protein TIGR04518; methyl-folate recycling enzymes (EC2.1.1.13 and EC1.5.1.54); methionine import proteins (EC2.1.1.10 and TIGR02314); folate retention and salvage enzymes (EC6.3.2.12, EC6.3.2.17); and enzymes needed for downstream folate-dependent biosynthetic pathways that can remain functional if folate is acquired through alternative mechanisms (EC6.3.4.3, EC3.5.4.9, EC2.1.2.2 for purine biosynthesis; and EC2.1.2.1 for purine and dTMP biosynthesis) (**Extended Data Fig. 2**). Genomes were hierarchically clustered (Ward’s method, Manhattan distance) based on binary presence/absence of these 15 gene functions. Silhouette scores were evaluated across k=2-20 (**Extended Data Fig. 3**). Mean silhouette width increased across the full range without reaching a clear inflection point, consistent with the binary, low-dimensional nature of the gene presence/absence matrix. k=12 was selected on the basis of parsimony and biological interpretability as the smallest number of clusters resolving functionally distinct folate pathway architectures.

Genome-level survival prediction under SP pressure was determined using a rule-based model. Each genome was first classified not susceptible (neither DHPS nor DHFR present), partially susceptible (one of DHPS or DHFR present), or fully susceptible (both present). Survival of susceptible genomes under SP pressure were assigned through ThyX presence or exogenous folate import.

Bacteria encoding ThyX possess a folate-independent alternative to the canonical ThyA-mediated dTMP synthesis route. Unlike ThyA, which consumes 5,10-methylene-THF and requires DHFR to regenerate the oxidized product, ThyX uses flavin adenine dinucleotide (FADH₂) as the reductant and does not deplete the folate pool. Bacteria encoding ThyX can therefore maintain DNA synthesis under antifolate pressure. Survival via folate importation to sustain folate-dependent DNA synthesis was predicted when TIGR04518 was present without EC1.5.1.54. If EC1.5.1.54 was also present, EC2.1.1.13 was required for a genome to be predicted to be enriched, under the working assumption that EC1.5.1.54 creates a folate sink unless balanced by EC2.1.1.13.

To evaluate whether folate-pathway architecture could help explain the direction of differential abundance in vivo, we mapped each differentially abundant taxon to the folate-pathway profile of its corresponding IMG genome(s). This framework is exploratory and hypothesis-generating: it characterizes the genomic potential for antifolate tolerance while acknowledging that *in vivo* survival reflects ecological complexity beyond gene presence alone. Per cluster, we report the percentage of genomes carrying ThyX or meeting criteria for folate import as a survival mechanism. We also compare per cluster the proportion of genomes belonging to SP-enriched versus SP-reduced taxa using a within-cluster binomial test against a 50:50 null hypothesis, with Benjamini-Hochberg correction applied across all 12 clusters.

### Linear regression to test the association between SVs and GWG

The relative abundance of each non-sparse SV was log-transformed and expressed relative to that SV’s geometric mean abundance across all samples. For each SV, a geometric mean was computed across all samples as g(SV)=exp(mean(ln(x))). Each sample’s relative abundance for that SV was then expressed as ln(xᵢ / g(SV)), where xᵢ is the relative abundance in sample i. Zero values were replaced with a pseudocount of ε=1×10⁻⁶ prior to transformation. This yields values centred at zero across samples for each SV, interpretable as the log fold-deviation of a sample’s abundance from the population geometric mean for that SV. Microbiome associations with GWG was evaluated with 3 approaches:

1. cumulative exposure during treatment summarized as area under the curve (AUC) across consecutive protocol visits using the trapezoid rule. AUC captures both the magnitude and persistence of microbiome changes across the treatment period, providing a more complete summary of cumulative microbial exposure than a single time point. Larger AUC reflects higher and/or more sustained CLR relative abundances over more visits/with increasing doses; gaps between visits (e.g. visits 1 and 3 available but not visit 2 were linearly bridged. In these linear regressions, a 1 AUC unit corresponds to maintaining +1 CLR (≈2.7X relative abundance) between 2 visits.
2. after ≥3 doses using the relative abundance observed in the sample collected at the final available study visit, only if it was after ≥3 doses. The final available visit provides a cross-sectional snapshot of microbiome composition after the full course of treatment, complementing the longitudinal AUC summary.
3. aggregated as multi-SV composite predictors per participant to quantify the overall microbiome contribution to GWG. This was chosen because the relevant biological signal can be an ecological shift across multiple co-varying taxa rather than the behavior of any single organism.

We generated: **1)** a single overall composite by summing all differentially abundant SVs that associated with GWG in approaches #i and #ii into one score per participant; **2)** two composites by summing the SVs deemed to be enriched after SP (vs. DP) into one score and the reduced SVs into a separate score; **3)** four granular composite predictors grouping SVs by SP effect (enriched *vs*. reduced relative to DP) and by whether that change in their relative abundance after SP (vs. DP) had a positive or negative GWG effect (↑ vs ↓). For comparability across composites, sum scores were standardized (z-scored) prior to modeling, so effect estimates reflect the change in GWG z-score per 1 standard deviation (SD) increase in the microbiome composite.

An exploratory of p<0.1 was applied across all regression models to maximize sensitivity for candidate identification at this screening stage; downstream mediation and birthweight analyses provide independent validation of the selected SVs.

#### Analytical population for GWG linear regressions

IPTp doses were administered at each study visit, but not all women contributed stool samples at every consecutive visit. The analytical population was restricted to women who received ≥3 doses of IPTp, the recommended number of doses by the WHO (77 of 79 women with follow-up microbiome data; 2 women received fewer than 3 doses and were excluded). Among these, women were further required to have both a GWG measurement and at least one stool sample collected after the third dose. For AUC analyses, they were also required to have a stool sample collected after the first or second dose to compute the trapezoidal integral. GWG was measured at the last study visit prior to delivery, which occurred within approximately one month of delivery. Because IPTp doses were given approximately 1 month apart, microbiome profiles during IPTp are expected to reflect changes in the interval following the prior dose.

### Microbiome-mediated pathway analyses

We hypothesized that SP-induced microbiome shifts causally mediate part of SP’s total positive effect on GWG and that these microbiome-mediated effects on GWG associate with higher birthweight (expressed continuously in grams). We addressed this hypothesis in two analytical segments with distinct inferential status.

**Segment 1 (SP → microbiome → GWG; causal mediation)** used a parallel two-mediator structural equation model in Stata 19 to estimate the extent to which treatment-associated microbiome shifts mediated the effect of SP on gestational weight gain, following established methodology for multi-segment causal pathways^57^. This segment leverages the randomized design to support a causal interpretation of the SP → microbiome → GWG pathway^58^. Missing data were handled using full-information maximum likelihood (FIML; method(mlmv)) with robust standard errors (vce(robust)). Indirect effects were estimated as products of path coefficients (a×b) for each mediator composite and summed for the combined indirect effect (nlcom). Causal identification rests on the sequential ignorability assumption^58^. In this setting, the first condition, ignorability of treatment assignment, is supported by randomization. The second condition, no unmeasured mediator–outcome confounding conditional on treatment and prespecified baseline covariates, is assumed and cannot be verified empirically, representing an inherent limitation of mediation analysis^59^. Because treatment was randomized, the treatment-to-mediator and treatment-to-outcome components benefit from randomization; however, causal interpretation of the indirect effects additionally requires standard mediation assumptions, including no unmeasured mediator–outcome confounding conditional on treatment and prespecified baseline covariates, and no treatment-induced confounders of the mediator–outcome relationship. Accordingly, mediation estimates were interpreted as causal under these assumptions rather than as assumption-free causal proof. In sensitivity analyses, we assessed robustness of the indirect effect to alternative covariate adjustment sets and to prespecified residual mediator–outcome covariance structures representing unmeasured confounding. SV composites were used as mediators.

**Segment 2 (microbiome-predicted GWG → birthweight; associative)** estimated the association between microbiome-driven GWG variation and birthweight separately within each drug group. Within each drug group among women with three or more doses, individual microbiome composite scores were used to predict GWG via ordinary least squares regression. The resulting microbiome-predicted GWG values (representing the portion of GWG explained by microbiome composition within each treatment group) were then used as the independent variable in separate drug-stratified linear regression models with birthweight in grams as the outcome. This segment is associative rather than causal: although microbiome composition was shifted by randomized treatment in Segment 1, Segment 2 does not involve randomization into microbiome states and therefore supports an associative rather than causal interpretation of the microbiome-predicted GWG → birthweight relationship. Findings from Segment 2 should be interpreted as hypothesis-generating, providing an initial characterization of whether microbiome-mediated GWG variation associates with fetal growth outcomes in this sample.

These two segments are reported and interpreted independently. Segment 1 estimated causal mediation effects for the SP → microbiome → GWG pathway under standard identification assumptions, including randomized treatment assignment and no unmeasured mediator–outcome confounding conditional on baseline covariates. Segment 2 provides associative evidence for whether the microbiome-driven component of GWG relates to birthweight. Together they characterize the plausibility of a full microbiome-mediated pathway from SP to fetal growth, while accurately reflecting the inferential limits of each analytical step.

**Covariates.** Segment 1 models were run with and without adjustment for covariates known to influence GWG: baseline socioeconomic status (expressed as wealth tertiles determined using a household asset-based wealth index derived from data collected on various common household items and dwelling characteristics); gravidity; age at enrollment; gestational age at enrollment; middle-upper-arm-circumference (MUAC); height (cm); body-mass index (BMI kg/m²); malaria positivity by PCR at enrollment. To avoid collinearity in multivariable models, one covariate was selected from each category of correlated determinants based on established clinical and biological relationships. BMI was selected over MUAC and height as the standard index of pre-pregnancy nutritional status in GWG models, given that GWG guidelines are stratified to pre-pregnancy BMI and no equivalent GWG guidelines exist relative to MUAC^60^.

Segment 2 birthweight models were run with and without adjustment for maternal MUAC and gestational length at delivery. Here, MUAC replaced BMI as the index of maternal nutritional status, as MUAC more sensitively reflects the functional protein-energy reserves available for fetal growth and has demonstrated superior predictive validity for birthweight in sub-Saharan African populations, where BMI-based classifications are attenuated by the co-occurrence of low adiposity and depleted lean tissue mass^61^. Gravidity was selected over chronological age as the more direct determinant of GWG in this population. Malaria infection by PCR at enrollment and socioeconomic status were retained as independent covariates.

### AI assistance disclaimer

Large language models were used to assist with development of custom R scripts and language editing of the manuscript text. All code outputs were reviewed, tested and verified by the authors.

## Data availability

Raw 16S rRNA gene amplicon sequencing reads have been deposited in the NCBI Sequence Read Archive under BioProject accession PRJNA1416942 and are currently under embargo pending publication. A de-identified minimal dataset sufficient to reproduce the reported downstream analyses is available to editors and reviewers during peer review via a private link and will be deposited publicly upon publication in the Carolina Digital Repository. This dataset includes the processed sequence variant count table, relative abundance tables, and the de-identified sample metadata used in the manuscript analyses. Source data underlying the main figures are available to editors and reviewers during peer review and will be provided in full with the Article upon publication. Additional individual-level metadata are not publicly available because of participant privacy and consent restrictions but may be made available through an appropriate controlled-access mechanism where permitted.

## Code Availability

All custom analysis workflows used in this study are available in full to editors and reviewers during peer review via a private Figshare link. Upon publication, the exact version of the code used for this study will be deposited in a DOI-minting repository and made publicly available.

## Disclaimer

The contents of this manuscript are solely the responsibility of the authors and do not necessarily represent the official views of the US Centers for Disease Control and Prevention or the US Department of Health and Human Services.

## Author Contributions

Conceptualization, funding acquisition, writing - review & editing, project administration: JRG, SRM. Investigation, data curation, formal analysis, methodology, validation, visualization, writing - original draft, writing - review & editing: AW. Conceptualization, funding acquisition, writing - review & editing, resources, methodology: IC. Investigation, project administration, writing - review & editing: JC, DPM. Investigation, project administration: EM. Investigation: MK. Investigation, data curation: SMP. Methodology, supervision, writing - review & editing: JJJ.

## Acknowledgements

This study and its findings are dedicated to the amazing scientist and mentor that made it all happen, Steven R Meshnick. Your wisdom and support will never be forgotten. We would like to sincerely extend our gratitude to the study participants, their families, and their communities. We are thankful to Dr. José Villar, the Principal Investigator of INTERGROWTH-21, for his valuable advice and best practices in estimating gestational weight gain for the present study.

