## Extended Data Fig.1 for "Beyond malaria prevention: sulfadoxine-pyrimethamine treatment in pregnancy selectively remodels the maternal gut microbiome to increase gestational weight gain and improve birthweight"

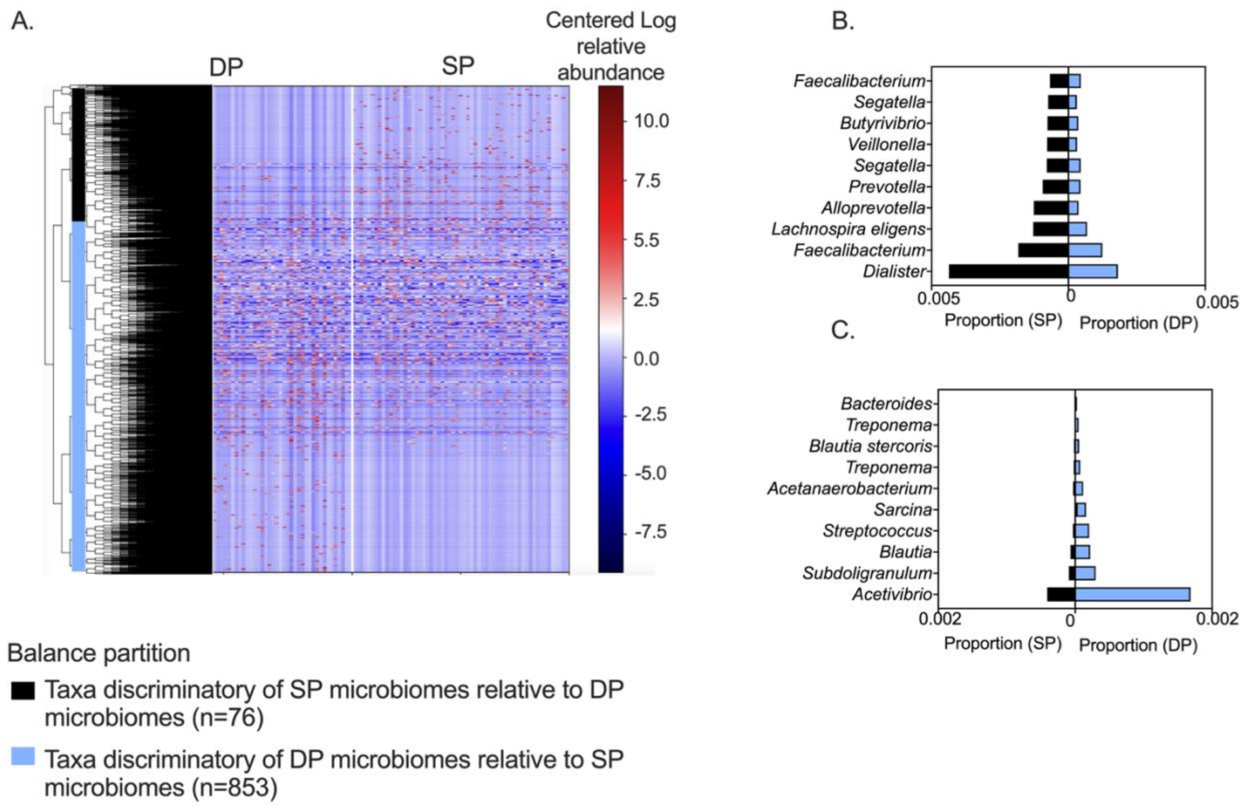


**Extended Data Fig 1. Exploratory identification of differentially abundant microbial sequence variants (SVs) by IPTp treatment group.**
(A) Heatmap of SV relative abundances (rows, centered log-ratio transformed) across all post-treatment samples (columns), grouped by IPTp drug: DP (left) and SP (right). SVs were hierarchically clustered using Gneiss (QIIME 2), which organizes microbes based on their co-variation across samples and identifies balances that differentiate groups. The top portion highlights the 76 SVs most discriminatory of SP-treated microbiomes, while the remainder (n=853) were more characteristic of DP-treated microbiomes. These discriminatory SVs were carried forward for focused comparison in Panels B and C.
(B) Top 10 SVs most characteristic of SP microbiomes, ranked by the largest directional loge difference in geometric mean abundance between SP and DP groups ($\log_e$ difference range=-0.7 to 1.5, corresponding to an approximate 2-fold to 4.5-fold difference). Bars indicate the relative contribution of each SV to the group of 76 SVs that were most discriminatory of SP microbiomes.
(C) Top 10 SVs most characteristic of DP-treated microbiomes, ranked by magnitude of loge difference in geometric mean abundance (loge difference range=-2.1 to 1.7, corresponding to an approximate 8-fold to 5.5-fold difference). These taxa are the strongest contributors to among the 853 that distinguished DP from SP microbial profiles. This exploratory analysis revealed strong, structured differences by treatment and informed the statistical validation in **Main Fig. 1**.

**Additional interpretation to Extended Data Fig. 1:** We employed the Gneiss tool implemented in QIIME2 as a first high-level exploratory step to determine if IPTp-SP *vs*. DP led to shifts in the relative abundance of microbes. To assess drug effects, we included only post-enrollment microbiomes (n=173). The Gneiss tree-based balance approach partitions sets of SVs that discriminate between groups, in our case IPTp drug group. A total of 929 microbial variants (sequence variants, SVs) were eligible to be compared across the after applying quality and sparsity filters (SVs with a minimum of 50 reads in a minimum of 5 samples). Different sets of microbes defined fecal microbiomes in the SP group relative to the DP group during the IPTp course. A set of 76 microbial variants over the course of the study were characteristic of microbiomes from SP mothers, while a distinct set of 853 SVs was enriched in DP microbiomes. Out of all the SVs that define the SP-associated microbial signature*, Faecalibacterium prausnitzii* SVs stands out in number and abundance relative. Higher abundance of *Streptococcus*, *Treponema*, *Bacteroides*, *Mogibacterium*, and lower abundance of *Ruminococcus*, *Eubacterium coprostanoligenes*, and *Bifidobacterium* contributed large fractions of the DP-specific signature. The taxa most discriminatory of SP microbiomes in the Gneiss analysis were largely recapitulated in the formal differential abundance analysis, providing independent cross-method support for the compositional differences identified between treatment arms.
