## Extended Data Fig.2 for "Beyond malaria prevention: sulfadoxine-pyrimethamine treatment in pregnancy selectively remodels the maternal gut microbiome to increase gestational weight gain and improve birthweight"

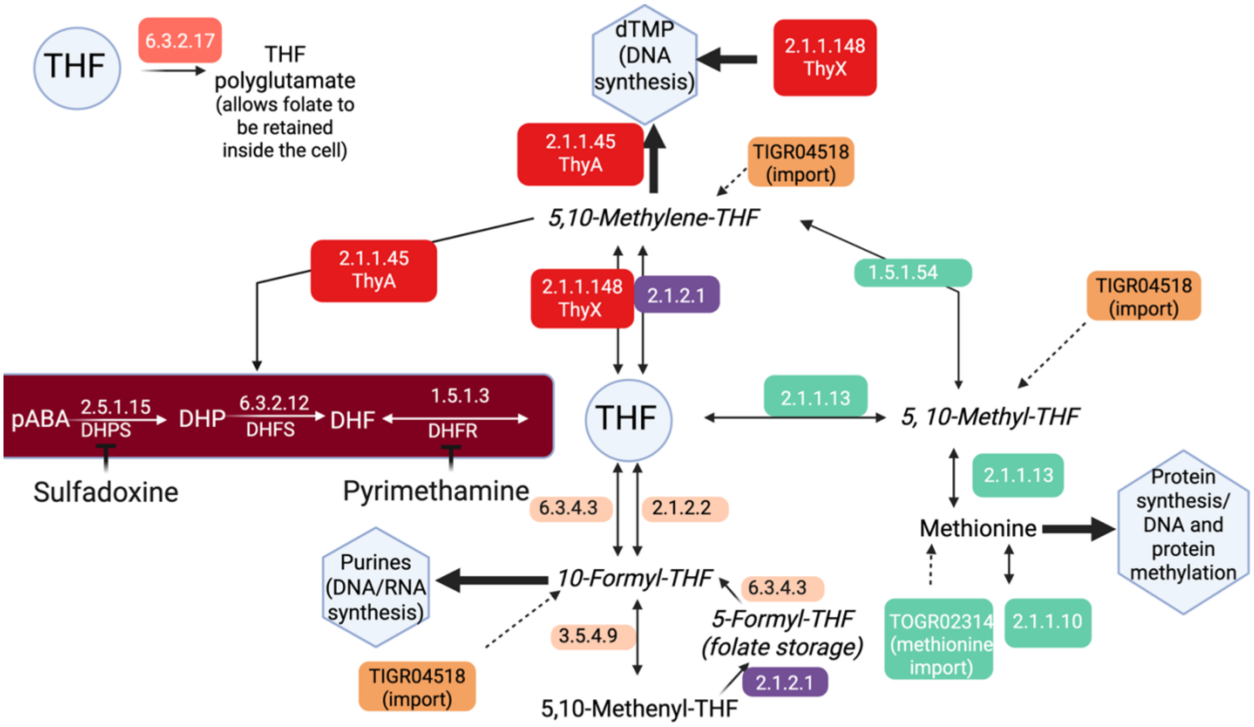


**Extended Data Fig. 2. Microbial folate and one-carbon metabolic pathways relevant for predicting survival under sulfadoxine-pyrimethamine (SP) antifolate pressure.** Sulfadoxine and pyrimethamine competitively inhibit dihydropteroate synthase (DHPS) and dihydrofolate reductase (DHFR) respectively, two sequential enzymes essential for microbial *de novo* synthesis of tetrahydrofolate (THF), the active folate cofactor required for the biosynthesis of nucleotides and amino acids. Bacteria that rely entirely on *de novo* THF synthesis are therefore predicted to be SP-susceptible. The Fig. illustrates the folate and one-carbon metabolic pathways that shape microbial survival capacity under SP pressure. Colored boxes indicate enzymes targeted by SP (red); enzymes whose presence confers potential survival capacity under SP pressure (green); and enzymes included in the gene panel for metabolic context but not used in the survival prediction rule (grey). Solid arrows indicate the canonical THF synthesis pathway; dashed arrows indicate alternative or bypass routes. Survival capacity under SP pressure was assessed through two independent mechanisms applicable to susceptible genomes: ThyX (EC2.1.1.148), which provides a folate-independent route to thymidylate (dTMP) synthesis by using $FADH_2$ as a reductant instead of 5,10-methylene-THF, thereby bypassing the DHFR requirement within the thymidylate branch; and qualifying folate import (TIGR04518), which restores the exogenous folate pool depleted by SP's inhibition of *de novo* synthesis. Folate import is counted as a survival mechanism only when EC1.5.1.54 is absent, or when both EC1.5.1.54 and EC2.1.1.13 are present; EC1.5.1.54 without EC2.1.1.13 is not counted because EC1.5.1.54 (methylenetetrahydrofolate reductase) irreversibly converts 5,10-methylene-THF to 5-methyl-THF, creating a folate sink that is only resolved when EC2.1.1.13 (methionine synthase) recycles 5-methyl-THF back to THF. Genomes lacking both SP targets (not susceptible) are predicted to survive by default. Methionine import (TIGR02314) and folate retention and salvage enzymes (EC6.3.2.12, EC6.3.2.17) are included in the gene panel for metabolic context but were not used in the survival prediction rule, as explained in Supplementary Note 2.
