## Extended Data Fig.3 for "Beyond malaria prevention: sulfadoxine-pyrimethamine treatment in pregnancy selectively remodels the maternal gut microbiome to increase gestational weight gain and improve birthweight"

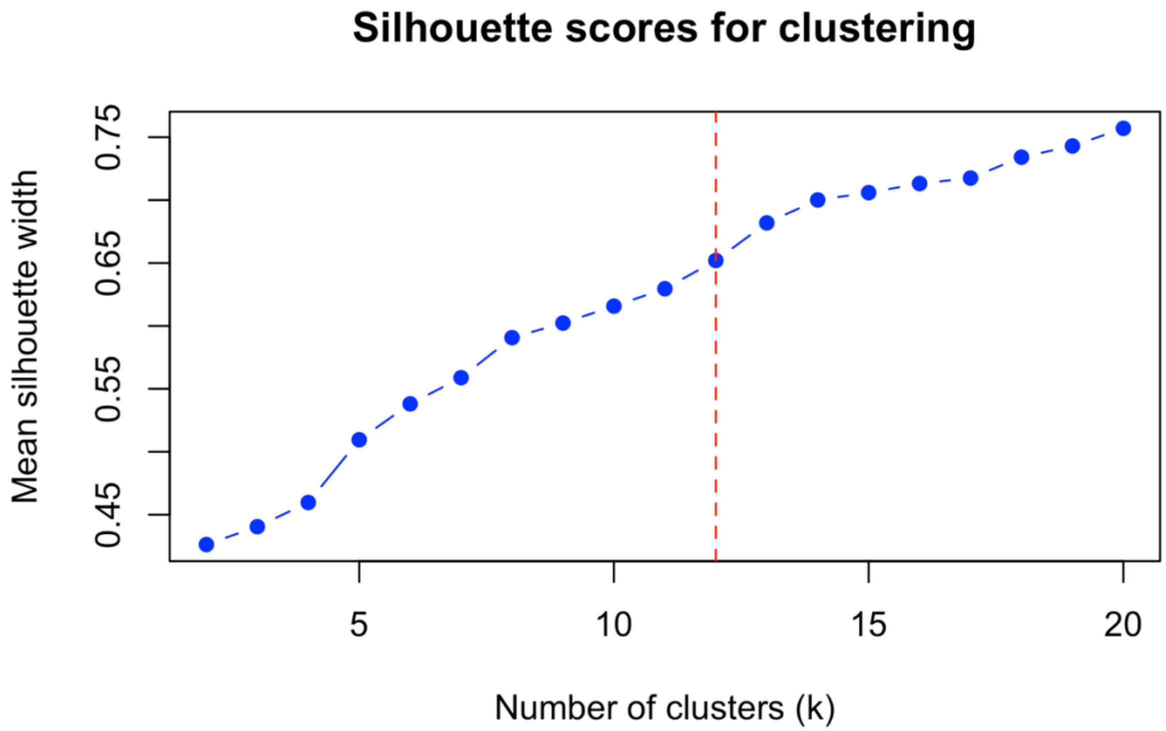


**Extended Data Fig. 3.** Silhouette score evaluation across k=2-20 for hierarchical clustering of 1,152 isolate genomes by folate pathway gene presence/absence (Ward's method, Manhattan distance). Mean silhouette width increases across the range evaluated, consistent with the binary, low-dimensional nature of the 15-gene presence/absence matrix in which finer partitioning continues to improve within-cluster homogeneity without producing a clear inflection point. In this setting, silhouette maximization does not identify a single optimal k; instead, k=12 (red dashed line) was selected on the basis of parsimony and biological interpretability.
