## Extended Data Fig.4 for "Beyond malaria prevention: sulfadoxine-pyrimethamine treatment in pregnancy selectively remodels the maternal gut microbiome to increase gestational weight gain and improve birthweight"

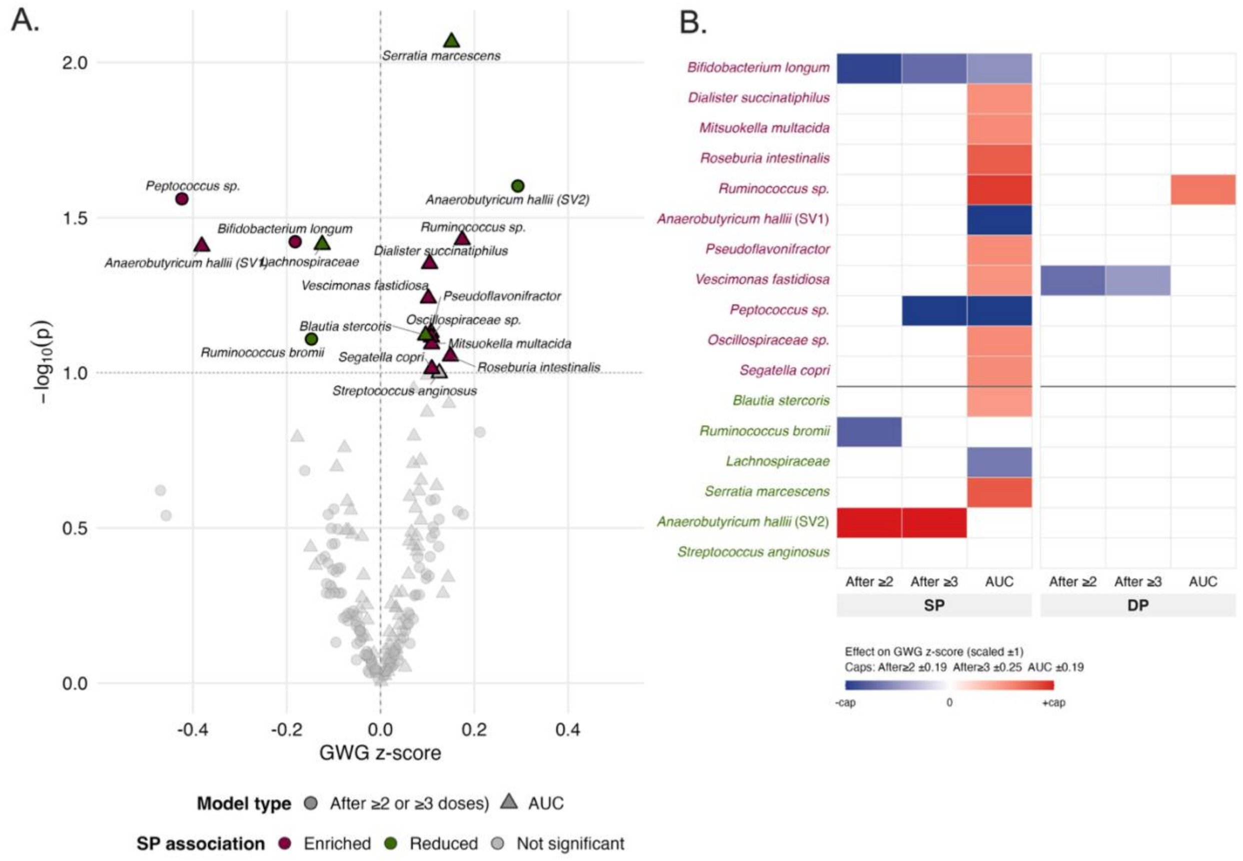


**Extended Data Fig. 4. Associations of gut microbiome sequence variants (SVs) with gestational weight gain (GWG) in SP-treated women.**

**(A)** Volcano plot showing associations between each of 84 differentially abundant SVs and GWG z-score in SP-treated women, across two analytic approaches: cross-sectional abundance using the stool samples most proximal to delivery, provided it was collected after ≥2 or ≥3 doses (circles) and cumulative area-under-the-curve (AUC) abundance across visits (triangles). Color indicates enriched (maroon) or reduced (green) status relative to DP in the differential abundance analysis, at the exploratory threshold of $p<0.1$; grey points did not reach this threshold. Black borders identify the 17 SVs carried forward to Fig. 3B. Unadjusted p-values; no multiple testing correction applied for this exploratory, hypothesis-generating analysis.

**(B)** Heatmap showing the direction and magnitude of associations between GWG z-score and each of the 17 selected SVs across model types (cross-sectionally using samples collected at ≥2, Raw ≥3, AUC) and drug groups (SP, DP). Rows show SVs ordered by hierarchical clustering within DA class (enriched above the horizontal line, reduced below). Only associations reaching $p<0.1$ are colored; non-significant cells are white. Effect sizes are scaled per model type to a unit range. Red indicates a positive association with GWG z-score; blue indicates a negative association. Taxon names are colored by DA class: maroon=enriched with SP vs. DP; green=reduced with SP vs. DP.
