## Extended Data Table 1 for "Beyond malaria prevention: sulfadoxine-pyrimethamine treatment in pregnancy selectively remodels the maternal gut microbiome to increase gestational weight gain and improve birthweight"

**Table 1. Baseline characteristics of the ancillary microbiome cohort by randomized group**

| Characteristic | Overall | IPTp-SP | IPTp-DP |
|---|---|---|---|
| Participants, n | 91 | 45 | 46 |
| **Maternal demographics** | | | |
| Age at enrollment, years | 20.9 (2.6) | 20.8 (2.5) | 20.9 (2.7) |
| **Gravidity category** | | | |
| 1 | 41 (45.1%) | 22 (48.9%) | 19 (41.3%) |
| 2 | 34 (37.4%) | 15 (33.3%) | 19 (41.3%) |
| 3 | 16 (17.6%) | 8 (17.8%) | 8 (17.4%) |
| **Wealth tertile** | | | |
| 1 | 43 (47.3%) | 19 (42.2%) | 24 (52.2%) |
| 2 | 25 (27.5%) | 13 (28.9%) | 12 (26.1%) |
| 3 | 23 (25.3%) | 13 (28.9%) | 10 (21.7%) |
| **Anthropometry** | | | |
| Weight at enrollment, kg | 56.3 (7.2) | 56.7 (7.6) | 55.9 (6.8) |
| MUAC at enrollment, cm | 25.8 (2.2) | 25.7 (2.1) | 26.0 (2.4) |
| Height, cm | 157.3 (5.6) | 157.8 (6.1) | 156.9 (5.0) |
| BMI at enrollment, $kg/m^2$ | 22.9 (2.7) | 22.9 (2.4) | 23.0 (2.9) |
| **Clinical** | | | |
| Malaria PCR positive at enrollment | 27 (29.7%) | 14 (31.1%) | 13 (28.3%) |

Values are mean (SD) or n (%). Percentages are calculated among participants with available data. No statistical tests were performed for baseline comparisons between randomized groups.
